# Assessment of 3D hemi-diaphragmatic motion via free-breathing dynamic MRI in pediatric thoracic insufficiency syndrome

**DOI:** 10.1101/2024.05.02.24306551

**Authors:** Mahdie Hosseini, Jayaram K. Udupa, You Hao, Yubing Tong, Caiyun Wu, Yusuf Akhtar, Mostafa Al-Noury, Shiva Shaghaghi, Joseph M. McDonough, David M. Biko, Samantha Gogel, Oscar H. Mayer, Patrick J. Cahill, Drew A. Torigian, Jason B. Anari

## Abstract

**Purpose:** Thoracic insufficiency syndrome (TIS) affects ventilatory function due to spinal and thoracic deformities limiting lung space and diaphragmatic motion. Corrective orthopedic surgery can be used to help normalize skeletal anatomy, restoring lung space and diaphragmatic motion. This study employs free-breathing dynamic MRI (dMRI) and quantifies the 3D motion of each hemi-diaphragm surface in normal and TIS patients, and evaluates effects of surgical intervention.

**Materials and Methods:** In a retrospective study of 149 pediatric patients with TIS and 190 healthy children, we constructed 4D images from free-breathing dMRI and manually delineated the diaphragm at end-expiration (EE) and end-inspiration (EI) time points. We automatically selected 25 points uniformly on each hemi-diaphragm surface, calculated their relative velocities between EE and EI, and derived mean velocities in 13 homologous regions for each hemi-diaphragm to provide measures of regional 3D hemi-diaphragm motion. T-testing was used to compare velocity changes before and after surgery, and to velocities in healthy controls.

**Results:** The posterior-central region of the right hemi-diaphragm exhibited the highest average velocity post-operatively. Posterior regions showed greater velocity changes after surgery in both right and left hemi-diaphragms. Surgical reduction of thoracic Cobb angle displayed a stronger correlation with changes in diaphragm velocity than reduction in lumbar Cobb angle. Following surgery, the anterior regions of the left hemi-diaphragm tended to approach a more normal state.

**Conclusion:** Quantification of regional motion of the 3D diaphragm surface in normal subjects and TIS patients via free-breathing dMRI is feasible. Derived measurements can be assessed in comparison to normal subjects to study TIS and the effects of surgery.

## Introduction

Thoracic insufficiency syndrome (TIS) is a clinical condition affecting lung growth and ventilatory function in pediatric patients due to spinal and thoracic deformities.^1^ The impairment in ventilatory function within TIS patients stems from various factors, including scoliosis, limited chest wall expansion, restricted lung volume, and compromised diaphragmatic motion.^2^ The alteration in chest structure also impacts the orientation of the diaphragm, reducing its capacity to generate force.^1^ If left untreated, scoliosis increases the risk of dependence on nasal oxygen or ventilator support, leading to respiratory failure and premature mortality.^2,3^ To address these challenges, reconstructive surgeries like the vertical expandable prosthetic titanium rib (VEPTR) procedure are often performed to promote lung growth when progressive deformities hinder normal development. However, there is currently no established method to evaluate diaphragmatic function following such procedures.^4^

The normal diaphragm does not move uniformly during active breathing, with the greatest movement typically occurring in the middle and posterior third regions.^5^ There is significant natural asymmetry between the morphological features and dynamics of normal right and left ventilatory volume components.^6^ The right hemi-diaphragm (RHD) dome is positioned higher than the left hemi-diaphragm (LHD) during spontaneous breathing due to different mechanical properties of the diaphragm, phrenic nerve innervation, and the effect of the surrounding anatomical structures.^7,8^ Recent studies have explored dynamic imaging methods that enable continuous assessment of the thorax and lungs throughout the respiratory cycle, without the need for breath holding.^1^ Dynamic magnetic resonance imaging (dMRI) is a noninvasive modality for quantitatively evaluating regional respiratory mechanics and assessing the efficacy of surgical treatment.^3^

In this study, we employed an advanced 4D reconstruction method to analyze the motion of each hemi-diaphragm surface via free-breathing thoracic dMRI. Our goal was to create dynamic 4D thoracic images, enhancing both visualization and quantification of abnormalities. Moreover, we sought to gain quantitative insights into diaphragmatic function, assess the impact of surgical intervention for TIS, and compare these findings with those from normal healthy controls. Such information is intended to contribute to future clinical decision-making.

## Materials and Methods

### Patients, subjects, and data sets

Our study utilized data from a research protocol approved by the Institutional Review Board at the Children’s Hospital of Philadelphia (CHOP) and the University of Pennsylvania, under the Health Insurance Portability and Accountability Act waiver. We retrospectively analyzed a dataset comprising 149 pediatric patients with diagnosed TIS who underwent our dMRI scanning sequence. Among these patients, 49 underwent VEPTR surgery with dMRI performed pre and post (paired subjects), and 100 (all VEPTR) were unpaired (70 pre-operation and 30 post-operation). Additionally, dMRI images were collected from a control group of 190 healthy children. Thoracic dMRI scans were obtained in a supine position for all subjects using the following dMRI scan protocol: 3T MRI scanner (Siemens Healthcare, Erlangen, Germany), true FISP imaging with a steady-state precession sequence; TR/TE = 3.82/1.91 msec; voxel size approximately 1x1x6 mm^3^; matrix = 320 x 320; bandwidth = 558 Hz; flip angle = 76°. The protocol involved obtaining 2D images at 30-40 contiguous sagittal locations across the chest, and for each location, 80 slices were acquired continuously at a rate of approximately 400 msec per slice during natural breathing. These free-breathing dMRI acquisitions were used to construct a 4D dMRI image, representing the dynamic thoracic movement over one respiratory cycle.^9^

### Diaphragm surface formation

A trained human tracer manually delineated the superior boundary of the diaphragm on each sagittal slice of the 4D constructed image at end-expiration (EE) and end-inspiration (EI) time points, covering the entire diaphragm from its anterior to the posterior aspects as visible on the dMRI sagittal slices, including areas adjacent to neighboring organs or tissue structures such as the heart, liver, or mediastinal fat (Figure 1). Subsequently, for both EE and EI time points in the cycle, we divided the diaphragm into the RHD and LHD using the midline sagittal slice and created hemi-surfaces separately for RHD and LHD from the respective segmentations. To assess the 3D movement of the diaphragm, we automatically selected 25 points uniformly on each hemi-diaphragm surface^10^, as illustrated in Figure 2.

**Figure 1.**
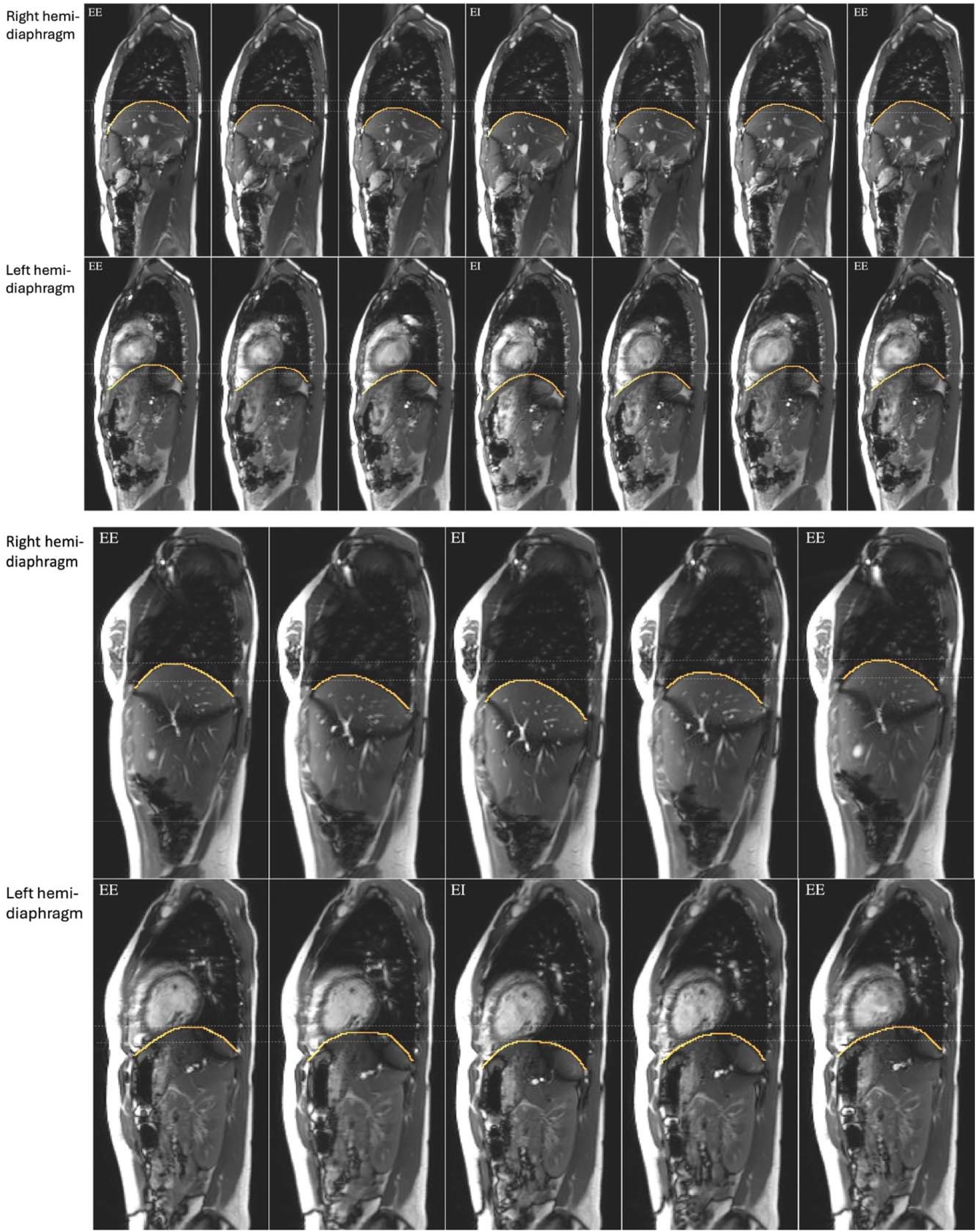
Sagittal slices from a 4D constructed image of a normal male (top two rows) and a normal female (bottom two rows) through the middle of the right hemi-diaphragm and left hemi-diaphragm are shown at different phases over one respiratory cycle. The segmented boundary is also displayed. EE: end-expiratory, EI: end-inspiratory.

**Figure 2.**
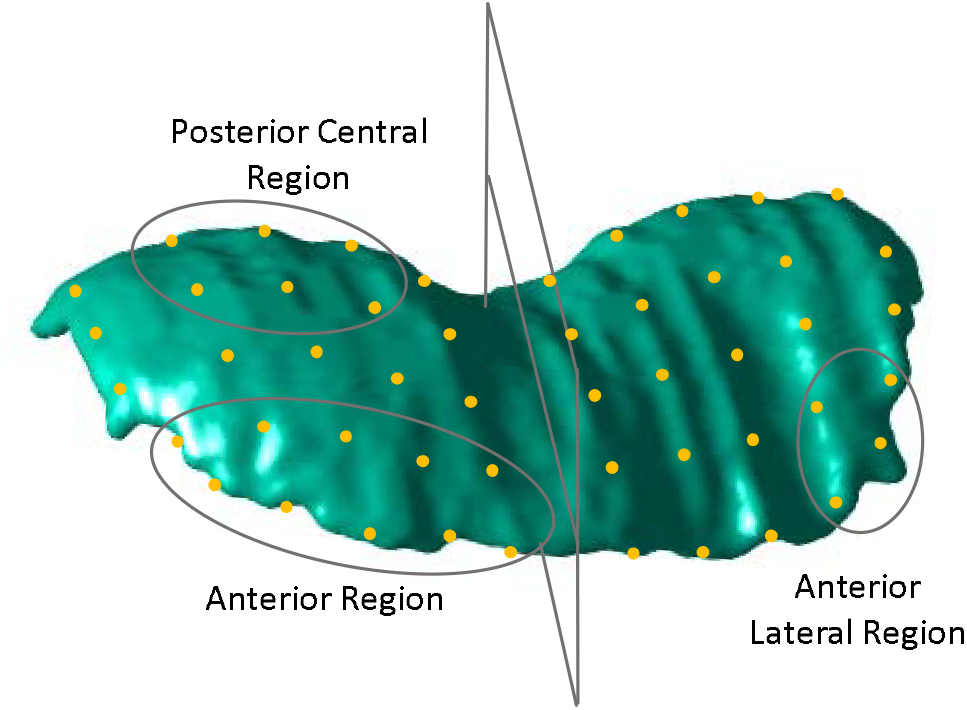
The diaphragm 3D surface is divided into left hemi-diaphragm (LHD) and right hemi-diaphragm (RHD) components using the mid-sagittal plane. On each hemi-diaphragm surface, 25 roughly equally distributed points are selected at each of the end-inspiration (EI) and end-expiration (EE) time points. The figure shows the diaphragm at the EE time point. For describing regional motion, several regions are defined as collections of points on each hemi-surface. Three exemplar regions are shown – Anterior region and Posterior Central region on the RHD and Anterior Lateral region on the LHD. The dMRI data set pertains to a healthy control subject.

### Velocity estimation

We calculated the velocity of each point on each hemi-diaphragm based on its superior-inferior displacement between the EE and EI time points (in units of mm/s).^10^ To estimate regional 3D motion, we organized the 25 points into 13 anatomic regions and calculated the average velocities within each region for each hemi-diaphragm surface. The velocity magnitude in the superior-inferior direction was preferred over displacement measurement as this approach normalizes displacements from subjects of different sizes, ensuring that the analysis is not skewed by variation in anatomical proportions. Moreover, since velocities are scalars measured in the superior-inferior direction, they can be averaged from multiple points to estimate regional velocity. The mean velocity for a region was calculated over all points within the region. The 13 regions were defined as follows: 1) Anterior Region (AR), 2) Posterior Region (PR), 3) Lateral Region (LR), 4) Medial Region (MR), 5) Central Region (CR), 6) Anterior-Lateral Region (ALR), 7) Anterior-Medial Region (AMR), 8) Anterior-Central Region (ACR), 9) Posterior-Lateral Region (PLR), 10) Posterior-Medial Region (PMR), 11) Posterior-Central Region (PCR), 12) Central-Lateral Region (CLR), and 13) Central-Medial Region (CMR). Note that there is overlap among some of the regions.^10^ For example, in RHD, two points are common between posterior central region and posterior lateral region (Figure 2).

### Statistical analyses

We categorized the 49 paired subjects into distinct subgroups based on Cobb angle of the spinal curve, a key metric for measuring the magnitude of spinal deformities, particularly in scoliosis, in order to minimize heterogeneity among subjects in terms of severity of the deformity. We refrained from differentiating between right or left-sided curves, as most subjects (32 out of 49) had their thoracic apex toward the right and lumbar apex toward the left. Furthermore, we were mainly interested in examining pre-to post-operation changes and comparing them with normal subjects; the handedness of curves will be considered in a separate paper for comprehensive analysis. Thoracic and lumbar Cobb angles were measured using frontal radiographs. Subgroup classification was determined by the alterations in the thoracic or lumbar Cobb angle following surgery as follows:

1. Subgroup where at least 1 Cobb angle decreased: This group included subjects whose thoracic or lumbar Cobb angle decreased after surgery.
2. Subgroup where both thoracic and lumbar Cobb angles decreased: Subjects in this group experienced a decrease in both thoracic and lumbar Cobb angles after surgery.
3. Subgroup where thoracic Cobb angle decreased: Subjects in this group exhibited a decrease solely in the thoracic Cobb angle after surgery.
4. Subgroup where lumbar Cobb angle decreased: This group comprised subjects who showed a decrease solely in the lumbar Cobb angle after surgery.

We performed regression analysis in healthy children to understand the motion trends of each region in relation to age. Since healthy children in our cohort are older than some of our TIS patients, we employed the regression functions to estimate the velocities for younger ages of the healthy children and used these extrapolated values to compare against velocities from patients. Then, we conducted t-testing to compare the motion of the RHD and LHD between homologous regions among TIS patients, TIS patients before and after surgery, and TIS patients and age-matched normal healthy controls. A p-value less than 0.05 was considered to denote statistical significance.

## Results

The demographic information for 149 TIS patients and 190 healthy children is summarized in Table 1.

**Table 1.** Demographic information of 149 thoracic insufficiency syndrome (TIS) subjects and 190 healthy children.

| Dataset | Pre-operation (n) | Post-operation (n) | Females (%) | Males (%) | Mean age (SD) | Age range (years) |
| --- | --- | --- | --- | --- | --- | --- |
| 190 healthy children | N/A | N/A | 98 (51.6%) | 92 (48.4%) | 11.9 (3.6) | 6.2-18.9 |
| 49 paired TIS patients | 49 | 49 | 22 (44.9%) | 27 (55.1%) | Pre-op: 3.5 (3.5) | 0.3-12.7 |
|  |  |  |  |  | Post-op: 5.9 (3.6) | 1.6-14.5 |
| 100 unpaired TIS patients | 70 | 30 | 46 (46%) | 54 (54%) | Pre-op: 4.9 (4.7) | 0.2-18.1 |
|  |  |  |  |  | Post-op: 8 (4.3) | 0.8-18.5 |

### Comparing diaphragmatic motion pre- and post-operatively

Table 2 presents the mean and standard deviation (SD) of average velocities (mm/s) for the RHD and LHD in the cohort of 49 paired TIS subjects, derived from dMRI for 13 regions. According to Table 2, the posterior-central region (PCR) of the RHD exhibited the highest average velocity post-operatively. Significant velocity changes were observed in 7 out of the 13 regions (PR, MR, CR, PLR, PMR, PCR, CMR) of the RHD after surgery. For the LHD, statistically significant differences were found in only 2 regions out of the 13 regions (PR and PMR) following surgery. These findings collectively highlight more substantial changes in the RHD, specifically in the posterior-medial-central regions compared to the anterior-lateral regions.

**Table 2.** Means and standard deviations (SD) of average velocities (mm/s) in 13 regions for right hemi-diaphragm (RHD) and left hemi-diaphragm (LHD) obtained from dMRI in a cohort of 49 TIS subjects, along with p-values of pre-operative to post-operative comparisons. AR: anterior region; PR: posterior region; LR: lateral region; MR: medial region; CR: central region; ALR: anterior-lateral region; AMR: anterior-medial region; ACR: anterior-central region; PLR: posterior-lateral region; PMR: posterior-medial region; PCR: posterior-central region; CLR: central-lateral region; CMR: central-medial region. Statistically significant differences (p < 0.05) are shown in bold font.

|  |  | Velocities |  |  |  |  |  |  |  |  |  |  |  |  |
| --- | --- | --- | --- | --- | --- | --- | --- | --- | --- | --- | --- | --- | --- | --- |
|  |  | AR | PR | LR | MR | CR | ALR | AMR | ACR | PLR | PMR | PCR | CLR | CMR |
| RHD | Mean pre-op (SD) | 3.55 (1.75) | 3.88 (1.88) | 4.47 (1.97) | 3.06 (2.27) | 4.47 (1.80) | 4.26 (2.62) | 2.70 (2.39) | 3.97 (2.27) | 4.45 (2.01) | 3.24 (2.95) | 4.14 (2.21) | 4.70 (1.99) | 3.36 (2.38) |
|  | Mean post-op (SD) | 4.68 (4.34) | 6.28 (5.61) | 5.93 (4.92) | 4.88 (4.74) | 6.38 (4.94) | 5.54 (5.78) | 3.68 (3.38) | 5.36 (5.19) | 6.23 (4.74) | 6.08 (7.53) | **7.07** (6.85) | 6.05 (5.05) | 4.89 (4.13) |
|  | P-Value | 0.073 | **0.004** | 0.059 | **0.010** | **0.010** | 0.146 | 0.065 | 0.062 | **0.018** | **0.010** | **0.003** | 0.092 | **0.019** |
| LHD | Mean pre-op (SD) | 2.74 (1.79) | 3.08 (2.26) | 3.01 (2.01) | 3.11 (1.79) | 3.85 (2.02) | 2.37 (2.10) | 3.07 (2.17) | 2.89 (2.03) | 3.37 (3.15) | 2.86 (2.28) | 3.33 (2.62) | 3.35 (1.96) | 3.41 (1.83) |
|  | Mean post-op (SD) | 3.46 (4.53) | 4.22 (3.58) | 3.79 (3.54) | 4.35 (4.19) | 4.61 (4.91) | 3.25 (3.48) | 4.04 (5.26) | 3.55 (5.13) | 3.93 (4.07) | 4.33 (3.94) | 4.23 (3.88) | 4.39 (4.14) | 4.88 (4.75) |
|  | P-Value | 0.283 | **0.043** | 0.132 | 0.061 | 0.281 | 0.095 | 0.233 | 0.382 | 0.375 | **0.021** | 0.161 | 0.078 | 0.056 |

After categorizing the subjects based on changes of the spinal Cobb angle due to surgery, we noticed more significant changes in the RHD compared to those in the LHD. Additionally, we observed that a reduction in thoracic Cobb angle was linked to a higher number of statistically significant differences in the velocity of both hemi-diaphragms. This implies that reductions in the thoracic Cobb angle exhibited a stronger correlation with changes in diaphragm velocity compared to reductions in the lumbar Cobb angle (Table 3). Note that both thoracic and lumbar Cobb angles were reversed in three subjects after the operation. However, none exceeded the normal range (<10 degrees).

**Table 3.**
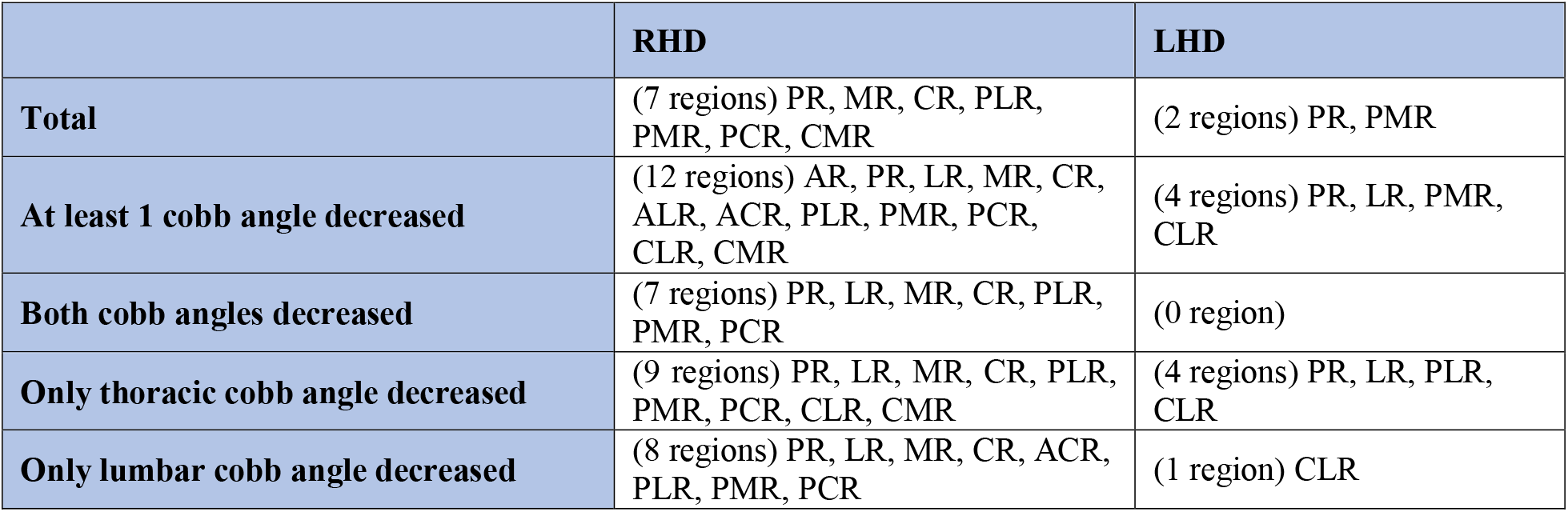
Statistically significant changes in diaphragm velocity for right hemi-diaphragm (RHD) and left hemi-diaphragm (LHD) in cohort of 49 paired TIS subjects based on changes in Cobb angle due to surgery. AR: anterior region; PR: posterior region; LR: lateral region; MR: medial region; CR: central region; ALR: anterior-lateral region; AMR: anterior-medial region; ACR: anterior-central region; PLR: posterior-lateral region; PMR: posterior-medial region; PCR: posterior-central region; CLR: centrallateral region; CMR: central-medial region.

### Comparing TIS diaphragm motion to normal group

Regression analysis performed on the cohort of 190 healthy children is displayed in Figure S1 in supplementary materials. Figure S1 reveals minimal alterations in diaphragm motion across a majority of 13 regions as age increased, both in the RHD and LHD, with some regions showing some increase in velocity with age (e.g., right PR, right PLR, and right PCR) and other showing decrease in velocity with age (e.g., left CR, left ACR, and left CLR). Table 4 demonstrates the p-values resulting from t-testing comparing diaphragm motion across 13 regions between 149 TIS subjects (both pre- and post-operatively) and the (extrapolated) values from the normal control group. ALR and ACR of the RHD and the anterior regions of LHD, including AR, ALR, AMR, and ACR, did not exhibit statistically significant differences post-operatively when compared to homologous regions in the normal control group. This suggests a trend wherein the anterior regions tended to approach normality after surgery while the posterior regions where the velocity is higher tended not to approach normality.

**Table 4.** p-values from t-testing, comparing motion of right hemi-diaphragm (RHD) and left hemidiaphragm (LHD) between 149 TIS subjects (both pre- and post-operatively) and normal controls for 13 regions. AR: anterior region; PR: posterior region; LR: lateral region; MR: medial region; CR: central region; ALR: anterior-lateral region; AMR: anterior-medial region; ACR: anterior-central region; PLR: posterior-lateral region; PMR: posterior-medial region; PCR: posterior-central region; CLR: centrallateral region; CMR: central-medial region. Statistically non-significant values (p ≥ 0.05) are shown in bold font.

|  |  | **AR** | **PR** | **LR** | **MR** | **CR** | **ALR** | **AMR** | **ACR** | **PLR** | **PMR** | **PCR** | **CLR** | **CMR** |
| --- | --- | --- | --- | --- | --- | --- | --- | --- | --- | --- | --- | --- | --- | --- |
| **Pre-op and control group** | **RHD** | 0.00 | 0.00 | 0.00 | 0.00 | 0.00 | 0.00 | 0.00 | 0.00 | 0.00 | 0.00 | 0.00 | 0.00 | 0.00 |
|  | **LHD** | 0.00 | 0.00 | 0.00 | 0.00 | 0.00 | 0.00 | 0.00 | 0.00 | 0.00 | 0.00 | 0.00 | 0.00 | 0.00 |
| **Post-op and control group** | **RHD** | 0.00 | 0.00 | 0.00 | 0.00 | 0.00 | **0.11** | 0.00 | 0.03 | 0.00 | 0.00 | 0.00 | 0.00 | 0.00 |
|  | **LHD** | **0.28** | 0.00 | 0.00 | 0.00 | 0.00 | 0.04 | **0.62** | **0.58** | 0.00 | 0.00 | 0.00 | 0.00 | 0.00 |

## Discussion

The diaphragm is the main breathing muscle and has significant importance in ventilatory function. The evaluation of diaphragm movement is essential for conditions that interfere with its function.^5^ Previous studies on pulmonary function have used varied techniques like pulmonary function testing, plethysmography, fluoroscopy, computed tomography (CT), or ultrasonography. However, these methods either fail to provide comprehensive information about diaphragmatic movements, especially 3-dimensional and regional, or come with practical drawbacks such as depending on patient cooperation or ionizing radiation exposure.^11,12^

Certain previous studies based on MRI have demonstrated that the 3D reconstruction method is accurate for measuring lung volumes and diaphragmatic components of the ventilatory pump.^6,13^ To our knowledge, this study is the first to investigate 3D diaphragm motion using free-breathing dMRI, specifically concerning pediatric ailments related to TIS. In this study, we applied dMRI to 149 pediatric patients with TIS and 190 normal children and assessed regional diaphragm motion and its changes following surgery. Unlike other currently available methods, this 4D dMRI method offers a practical noninvasive tool for the quantitative evaluation of the diaphragmatic regions of the ventilatory pump, making it suitable for routine clinical implementation.

Our findings generally suggest a notable increase in the motion of posterior diaphragm regions after surgery, whereas the anterior regions tend to normalize following the VEPTR procedure for pediatric TIS. Our previous study on the effect of scoliotic spinal curves in TIS using dMRI showed a qualitative postoperative increase in the chest wall and diaphragmatic excursion.^4^ Additionally, Yang X. et al.^5^ conducted a study on diaphragmatic motion in normal individuals using thoracic ratios and free-breathing dMRI. Their findings demonstrated a heightened prominence in the motions of the diaphragm apex and posterior diaphragm compared to the anterior diaphragm, supporting our observed trends. However, a study by Chu W. et al.^2^ utilizing breath-hold dMRI assessments found no difference in diaphragmatic motion between adolescent patients with idiopathic scoliosis and controls. This may indicate the importance of using a free-breathing method to accurately assess diaphragmatic motion under natural conditions.

The primary limitation of our study is the relatively small sample size of the patient cohort, especially for patients with both pre- and post-operative dMRI. However, our study used a larger sample size compared to currently available data. Another limitation is that manual segmentations of the dMRI scans were used in our study. As such, challenges in evaluating the location of the diaphragm contour and in identifying diaphragmatic attachments could be a factor for measurement variability. We are in the process of automating this process.^14^

In conclusion, this study introduces an advanced image analysis method for the quantification of the regional motion of the 3D diaphragm surface in pediatric TIS patients and normal healthy controls in a comparative fashion via free-breathing dMRI. The technique involved the generation of 4D images during free-breathing and meticulous tracing of the diaphragm in sagittal dynamic MRI images. Using this method, the regional motion of the two 3D hemi-diaphragm surfaces can be assessed quantitatively to determine the impact of TIS upon ventilatory function and the effects related to corrective surgery, generating new insights into the function of the diaphragm non-invasively, in its natural and free-breathing form, and 3-dimensionally. The quantitative information derived from dynamic MRI may potentially lead to improved treatment planning and patient outcomes.

## Data Availability

All data produced in the present study are available upon reasonable request to the authors.

## Acknowledgments

This work is supported by a grant R01HL150147 from the National Institutes of Health.

