## Supplementary material for "Assessment of 3D hemi-diaphragmatic motion via free-breathing dynamic MRI in pediatric thoracic insufficiency syndrome": Figure S1

**Supplementary materials**

**Figure S1.** Regression diagrams for right hemi-diaphragm (RHD) (left column) and left hemi-diaphragm (LHD) (right column) for 13 regions. AR: anterior region; PR: posterior region; LR: lateral region; MR: medial region; CR: central region; ALR: anterior-lateral region; AMR: anterior-medial region; ACR: anterior-central region; PLR: posterior-lateral region; PMR: posterior-medial region; PCR: posterior-central region; CLR: central-lateral region; CMR: central-medial region.

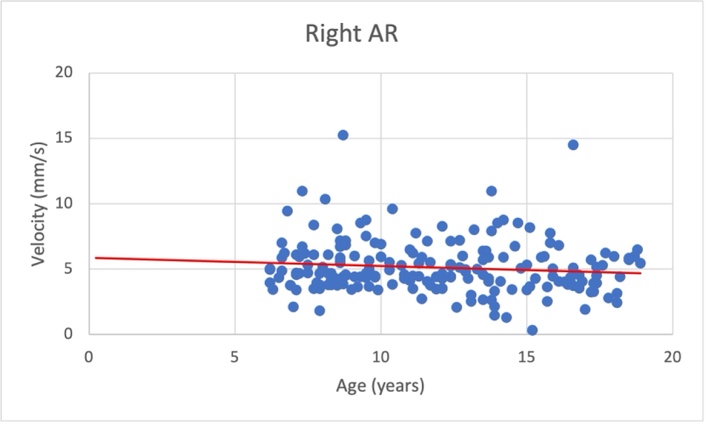

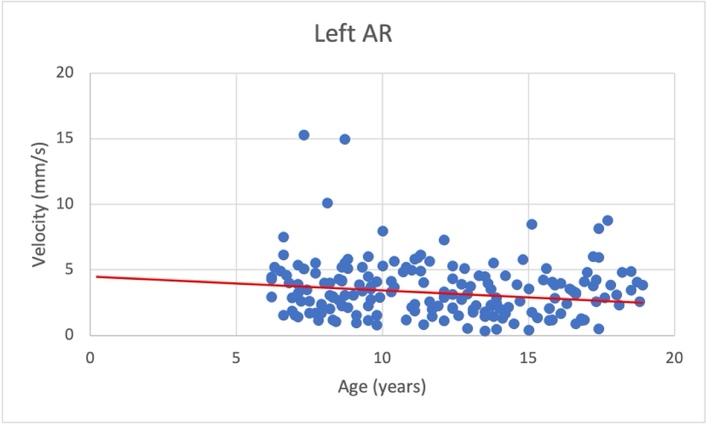

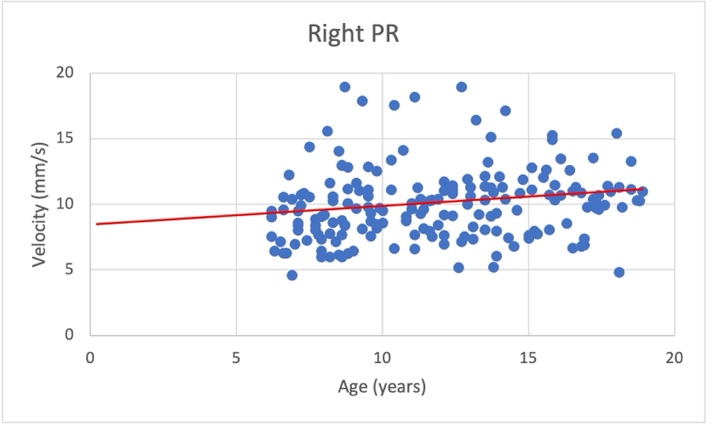

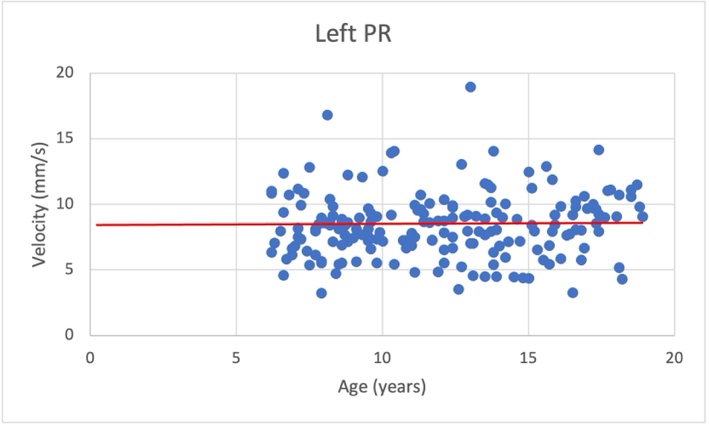

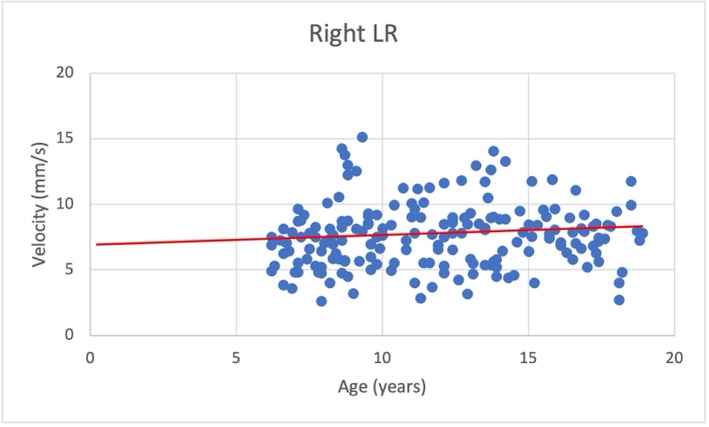

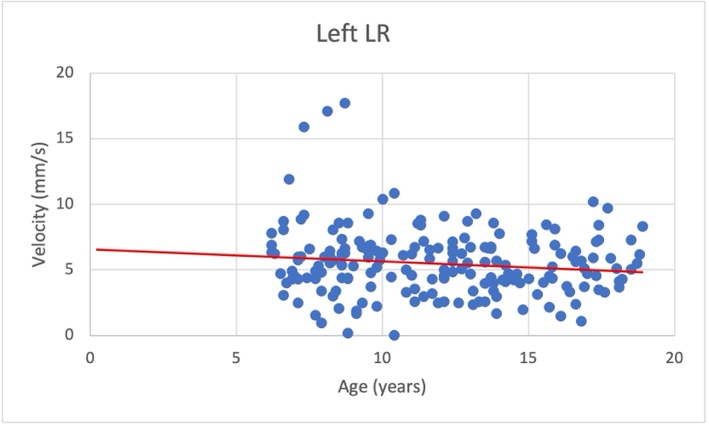

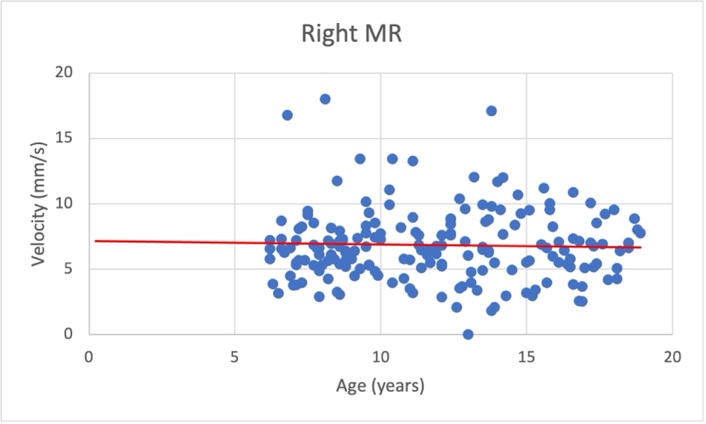

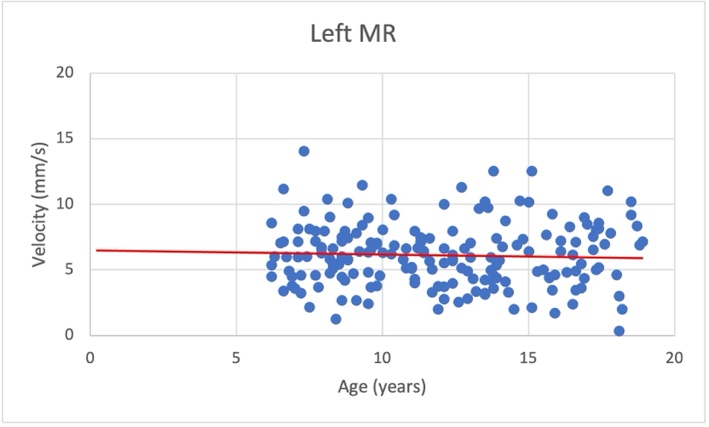

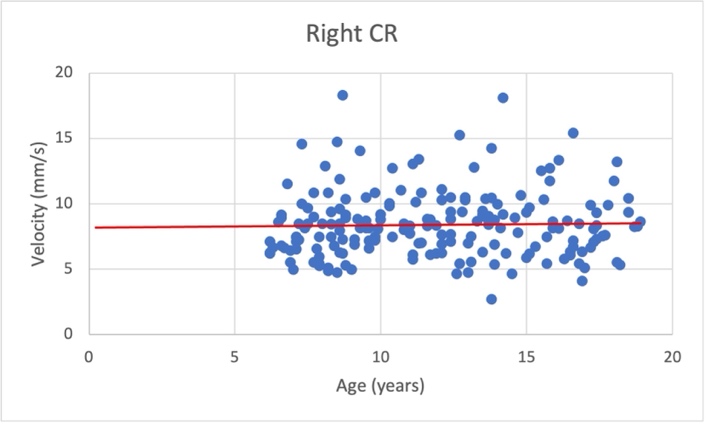

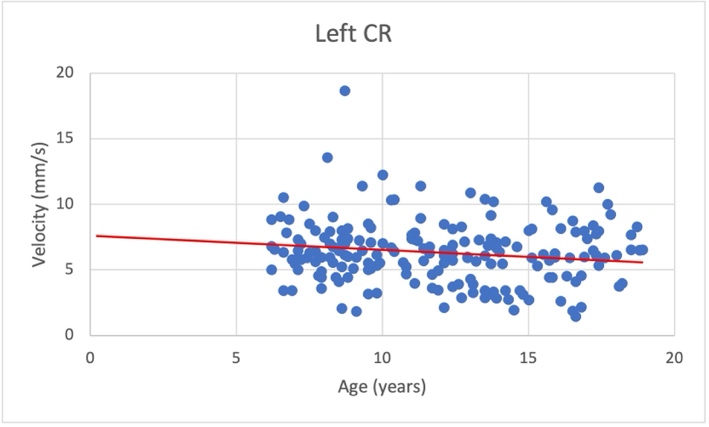

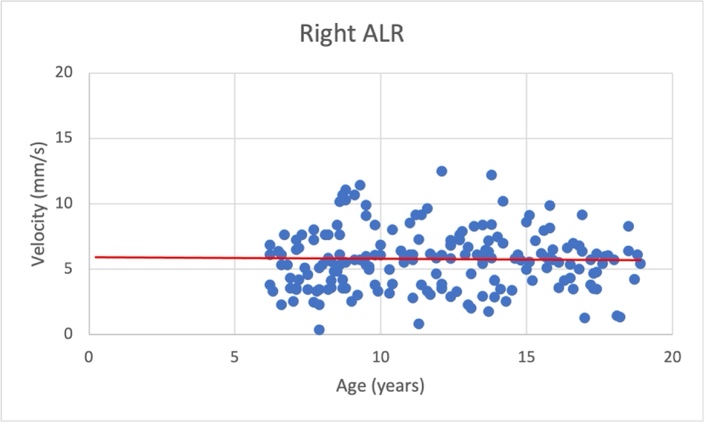

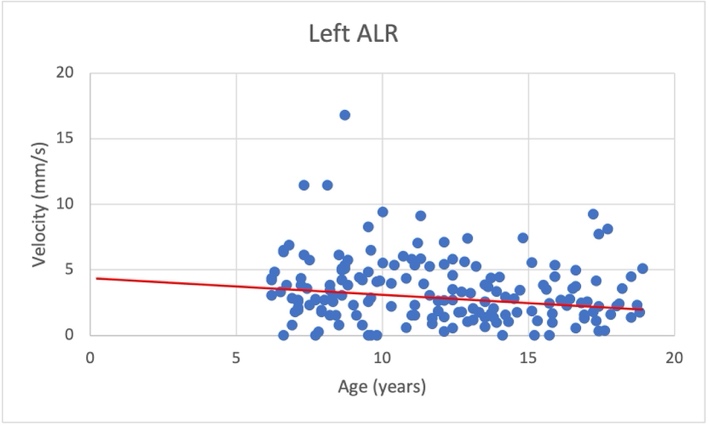

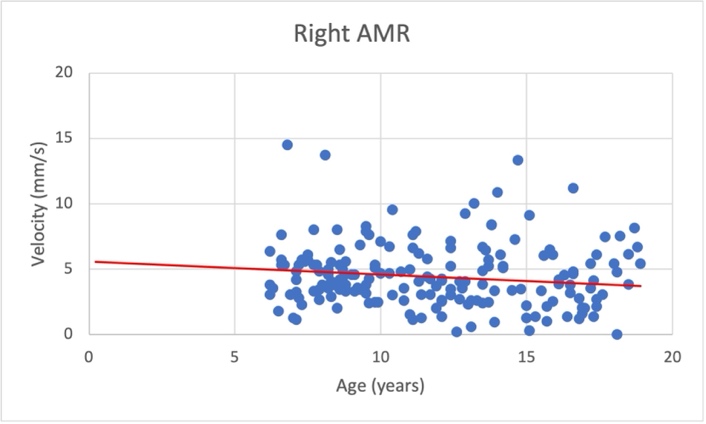

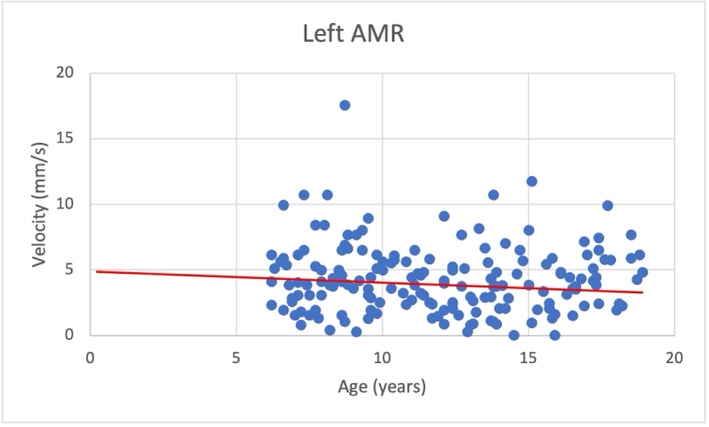

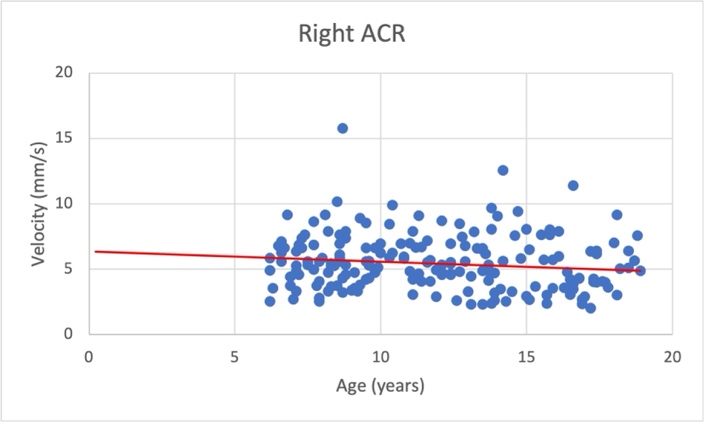

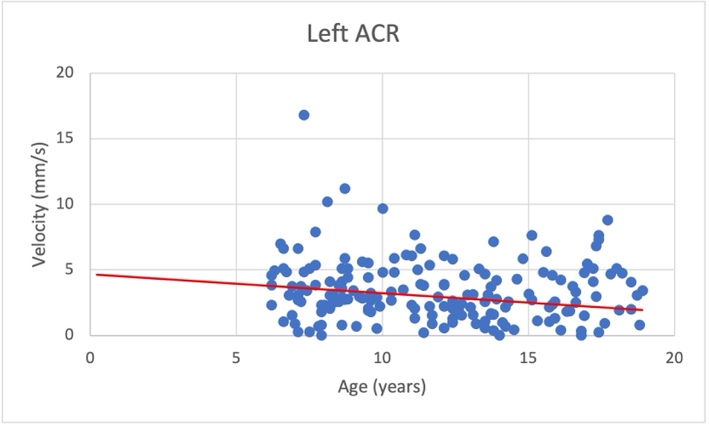

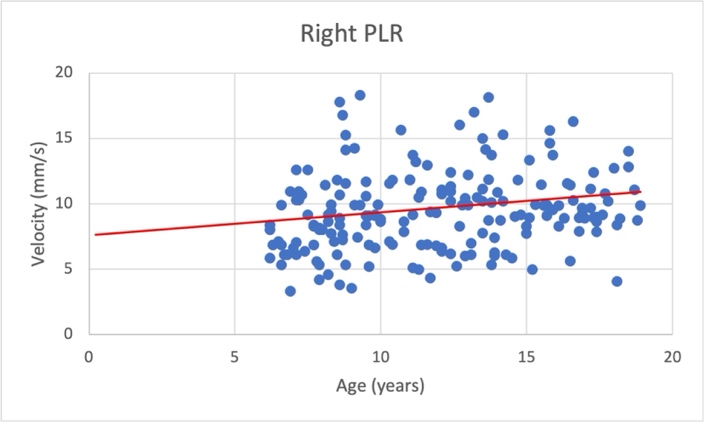

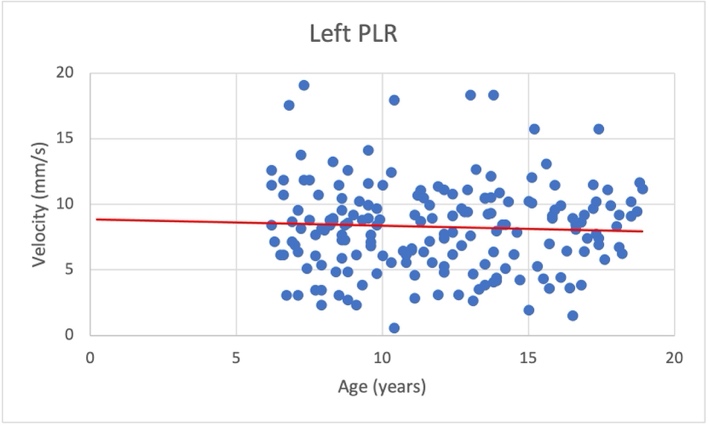

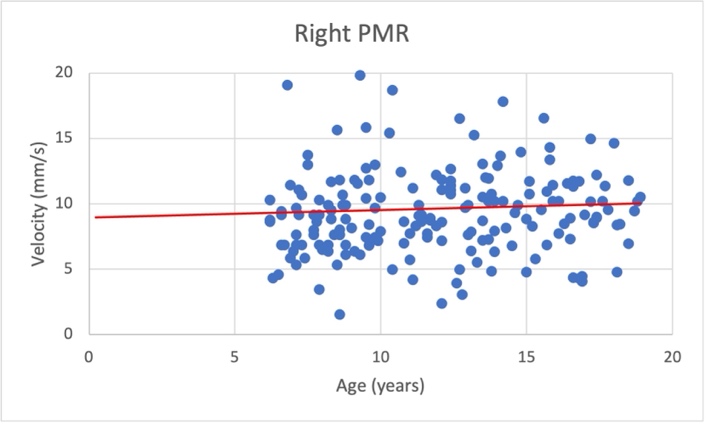

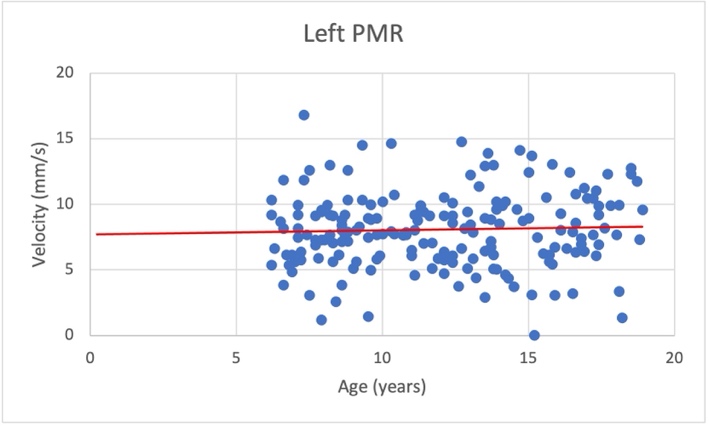

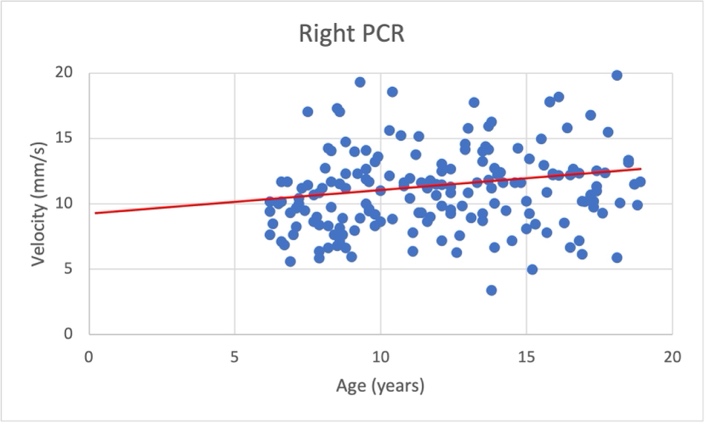

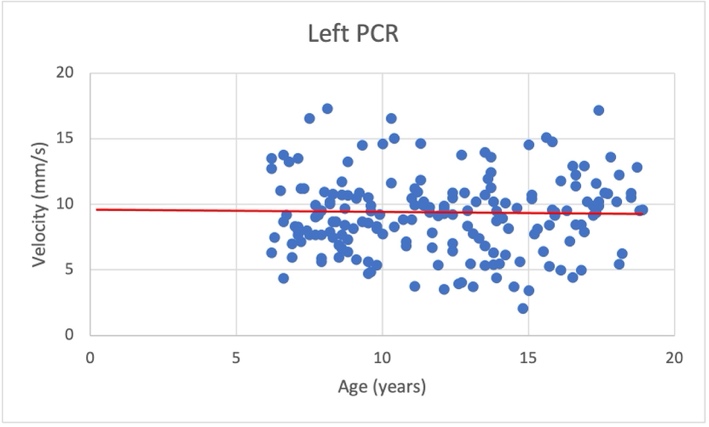

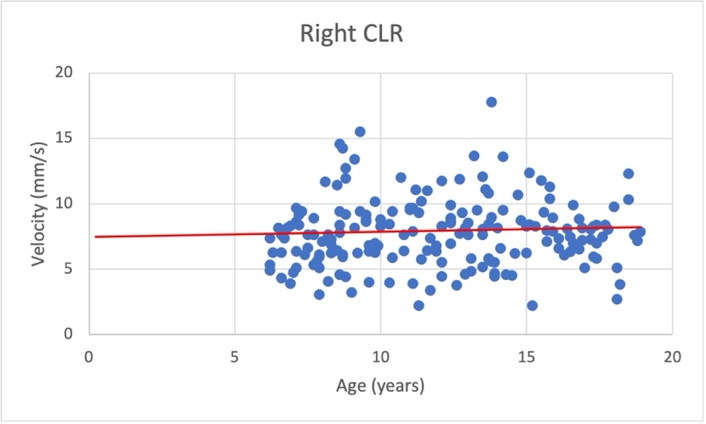

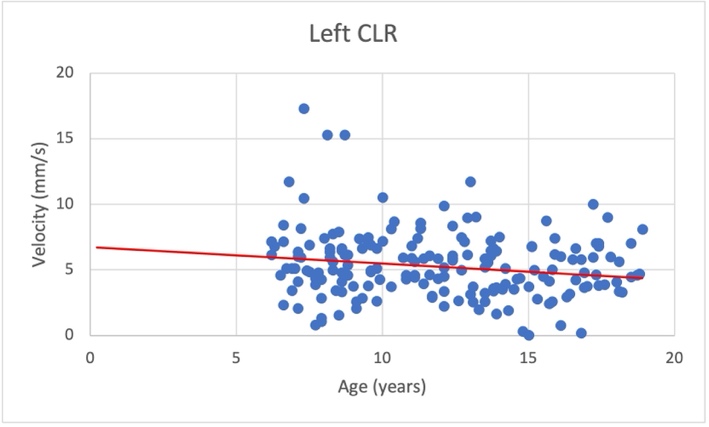

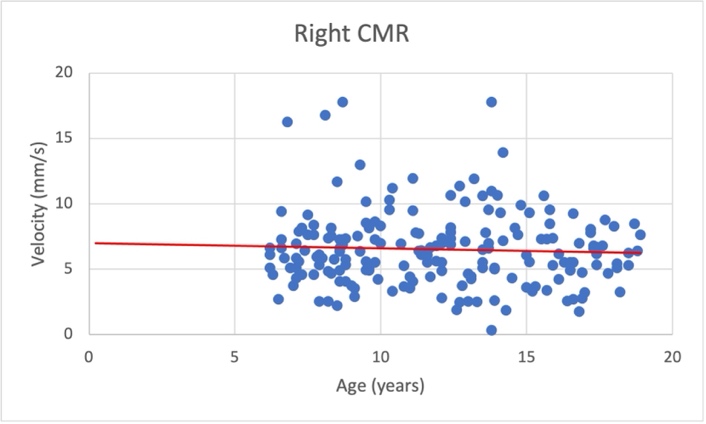

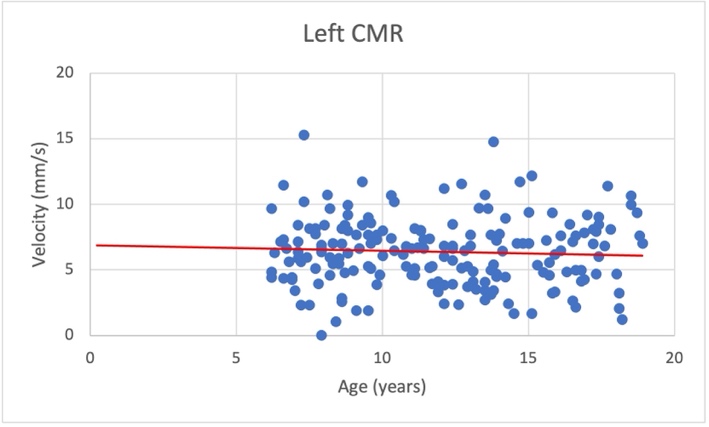
